# Protocol for a scoping review investigating success in research capacity building for nurses, midwives and allied health professionals

**DOI:** 10.1101/2025.03.07.25323570

**Authors:** Colin Hamilton, Natalie Pike, Lok Yiu Wong, Kieran Lock, Emma Jones, Gabrielle Deora, Graham Martin, Joanne McPeake

## Abstract

**Objective:** To identify and describe how success is currently conceptualised in research capacity building in nurses, midwives and allied health professionals in the UK.

**Introduction:** Having a research active healthcare workforce is associated with improved patient outcomes as well as staff retention. It is therefore seen as a key target for many healthcare organisations. Nurses, Midwives and Allied Health Professionals form the largest group of healthcare professionals but are traditionally less involved in research than medically trained staff. A variety of schemes have aimed to address this through so called “research capacity building” activities but an understanding of what constitutes success is needed to aid development of future interventions.

**Inclusion criteria:** Participants - Any or all of Nurses, Midwives or Allied Health Professionals

Concept-Definition of success or description of aims of activities aimed at research capacity building.

Context-Within in the UK.

**Methods:** Content from peer reviewed journals will be searched for in: Embase, CINAHL, MEDLINE, AMED, BNI and EMCARE Web of Science Core Collection.

Grey Literature will be searched for in Google and Overton as well as key websites of organisations that work in developing research capacity. Website searches will include National Institute for Health and Care Research, all charities that form the Association of Medical Research Charities as of search date and the websites for the recognised professional bodies for Nurses, Midwives and Allied Health Professionals.

Screening of titles and abstracts then full text will be undertaken by one person with 20% cross checked by a second reviewer. Data extraction will use a bespoke data extraction tool and will be undertaken by one person, with 20% cross checked with a second reviewer. A narrative synthesis and qualitative content analysis will be used to synthesise the data.

**Strengths and limitations of this study:**

- A wide range of information sources will be reviewed.
- A comprehensive search strategy has been developed in coordination with an experienced librarian.
- The project will focus on the UK; consequently applicability to other contexts will be limited.

## Introduction

Being a research active organisation has been associated with improved patient outcomes (1) and staff retention (2) as well as facilitating clinical developments (3). In the UK, clinical research is seen as a key element of the economy (4) as well as a way of managing challenges associated with aging populations amongst other issues (3). Engagement in research is therefore seen as a key goal for staff in the National Health Service (NHS) (5). There are significant barriers and challenges to engagement in research, however, which are well investigated and exist across many professions (6).

Nurses, Midwives and Allied Health Professionals (NMAHPs) form the majority of the patient-facing workforce in the NHS; however their engagement in research activities is low in comparison with medically trained staff (7). To address this many organisations have instigated the use of so-called “research capacity building” (RCB) exercises (8). These activities range from resources, webinars or single day courses, to fellowships and career paths (9). Significant time and money are invested in these activities which aim to produce outputs in the form of publications, further fellowships and active researchers (10). What constitutes “best practice” in research capacity building, however, is yet to be defined, and the evidence base is embryonic (8).

Part of the challenge seen is the lack of clarity on what constitutes success. Cooke et al (8) describe the aim of such activities as “doing more research, better”. However, this only appears to cover a small aspect of RCB and it does not provide a measure of what “doing more research” or what “better” is. When looking at something as complex as RCB the opinions of various stakeholders, including providers of RCB activities, funders, NMAHP consumers of these activities and healthcare managers, need to be considered, as their opinions on what success is are likely to vary depending on their own priorities.

An understanding of the range of criteria for success in RCB activities among the breadth of stakeholder groups is therefore required. While some clarity is likely to be produced in the published academic literature this is likely only a part of the picture and is limited in scope. As the RCB landscape is fragmented, with multiple funders and stakeholders, the grey literature is likely to be a significant source of understanding. The nebulous nature of RCB activities also calls for a flexible approach to understanding the current knowledge base.

Scoping reviews are advocated when attempting to clarify nebulous or multifaceted concepts of this kind (11). Scoping reviews do not attempt to appraise evidence or to provide any sort of hierarchy, which is appropriate when dealing with a concept like success in RCB where differing understandings can be equally valid. By building search strategies around population, concept and context, and reviewing a wide variety of literature sources, an overall understanding of the key elements can be built (11).

This project will undertake a scoping review to answer the question “How is success currently conceptualised in research capacity building in nurses, midwives and allied health professionals in the UK?”. A preliminary search of MEDLINE, the Cochrane Database of Systematic Reviews and Prospero was conducted and has identified no current systematic reviews or scoping reviews on this topic.

### Review aim

To identify and describe how success is currently conceptualised in research capacity building in nurses, midwives and allied health professionals in the UK.

### Review question

How is success currently conceptualised in research capacity building in nurses, midwives and allied health professionals in the UK?

### Inclusion criteria

#### Participants

Populations described should include one or all of Nurses, Midwives and the 14 Allied Health Professions.

#### Concept

Research capacity building or development covers a broad range of activities at the individual and organisational level (12). A generally accepted definition is that described by Trostle, of “a process of individual and institutional development which leads to higher levels of skills and greater ability to perform useful research”(13). Activities can include education, funding, advocacy and development of policy amongst others. In this context we will explore definitions of success or stated aims of activities which:

- Conform to Trostle’s definition given above
- Are described as research capacity building or development or similar

#### Context

The RCB landscape within the UK is unique but shares concepts with other countries. For literature in peer reviewed journals any healthcare context in the UK will be included. Concept papers or reviews may not state the context and will therefore be included if they do not exclude the UK. Grey literature which does not aim to be universally generalisable, including documents such as funders’ reports will be included if they relate to the UK.

#### Types of sources

This scoping review will consider articles published in peer reviewed journals of any sort including, but not limited to, conference abstracts, reviews, opinion pieces or primary research. It will also incorporate grey literature including funders’ reports, professional bodies and policy documents.

## Methods

The proposed scoping review will be conducted in accordance with the Joanna Briggs Institute methodology for scoping reviews(14).

### Search strategy

The search strategy will aim to identify both published and unpublished articles. A three-step search strategy will be utilized in this review. First, a limited initial search of MEDLINE (PubMed) and CINAHL (EBSCO) has been undertaken which identified 3 key papers (8,9,15). Key words in titles and abstracts as well as index terms and review search terms were used to develop a search strategy. Search strategies, where available, were screened for further terms. This search strategy was adapted for each database or information source and checked by a librarian. After implementing this search strategy as outlined, reference lists of included articles will be reviewed for possible further appropriate papers.

A full search strategy for sources can be found in Appendix 1.

#### Information sources will include

##### Published Literature

Through OVID: Embase, MEDLINE. Through EBSCO: CINAHL, EMCARE, BNI, Web of Science Core Collection.

##### Grey Literature

Google (first 100 results of search) and Overton.

##### Stakeholder documents and websites

National Institute for Health and Care Research

All charities that form the Association of Medical Research Charities as of search date.

The websites for the recognised professional bodies for Nurses, Midwives and AHPs.

### Study/Source of evidence selection

Following the search, all identified information sources will be collated and uploaded into the Rayyan review management software (www.rayyan.ai) and duplicates removed by reviewers. Titles and abstracts will then be screened by one reviewer with an independent reviewer checking a random 20% sample for assessment against the inclusion criteria. Any disagreements that arise between the reviewers will be resolved through discussion; if no resolution is possible then a third reviewer will arbitrate. Numbers of disagreements will be recorded.

The full text of selected items will be assessed against the inclusion criteria by one reviewer with a random 20% checked by a second independent reviewer. Reasons for exclusion will be recorded. Any disagreements that arise between the reviewers will be resolved through discussion; if no resolution is possible then a third reviewer will arbitrate. Numbers of disagreements will be recorded. The results of the search and the study inclusion process will be reported and presented in a PRISMA flow diagram.

### Data extraction

Data will be extracted from included papers by one reviewer using a bespoke data extraction tool (Appendix 2) developed for this review. A random 20% of entries will be checked by a second reviewer. The data extracted will include: title; year of publication; participants; context; study methods (if appropriate); author or publishing body (if appropriate); and description of aims or success.

The draft data extraction form was piloted on 2 papers that were found in the initial limited search. The data extraction tool may be modified during the review if needed from each included source. Modifications will be recorded and reported. Authors will be contacted for missing information or clarification if appropriate.

### Data analysis and presentation

In the first instance data will be presented in tabular form with key elements of each included paper shown. This will include (but not be limited to) author, date, type of paper (journal article, policy document, conference abstract etc), population and aim/description of success. A narrative summary will be produced to discuss the results and link them to the review objective.

It is presumed that aims of programmes or descriptions of success will have commonalities. An inductive qualitative content analysis will be undertaken to understand any commonalities (16). Two authors will gain deep familiarity with the data by reading and re-reading sources, then perform open coding. Following this, the two reviewers will meet to build a coding framework. Data will be extracted and organised using this framework. The coding framework will be revised, as necessary, to develop over-arching categories that address the review question and objectives.

## Patient and Public Involvement Statement

No patient or public involvement will be part of this project.

## Supporting information

Appendix 2 Data extraction tool

Appendix 1 Search Strategy

## Data Availability

All data produced in the present work are contained in the manuscript

## Acknowledgements

This review will form part of a PhD for CH.

## Author contributions

CH will undertake screening, data extraction, data analysis and write up. JM and GM will provide arbitration in any disagreements of inclusion or exclusion of information sources, support with conceptualising any coding framework and provide input into producing the initial draft and final draft of the review. AM will provide arbitration in any disagreements of inclusion or exclusion of information sources. NP, KL, EJ and LYW will undertake secondary reviewer of screening, data extraction, inductive content analysis and write up.

## Appendices

Appendix I: Search strategy

Appendix II: Data extraction instrument

