## Appendix 1 Search Strategy for "Protocol for a scoping review investigating success in research capacity building for nurses, midwives and allied health professionals"

**Search strategy- Medline via OVID**

**Population:**

**1**	("Nurs*").ti,ab,kw,kf. **OR** exp Nurses/ or exp Nursing/ or exp Nursing Staff/ or exp Nursing, Practical/ **OR** ("Midwives" or "Midwif*").ti,ab,kw,kf. **OR** exp Midwifery/ **OR (**"Allied Health* " or "AHP*" or "Professional* allied to health*" or "Art Therap*" or "dietitian*" or "dietetic*" or "Drama therap*" or "Music Therap*" or "Occupational Therap*" or "OT**" or "Operating Department Practitioner*" or "Operating room technician*" or "ODP" or "Orthoptist*" or "Orthoptic*" or "Osteopath*" or "Paramedic*" or "Podiatrist*" or "Podiatry" or "Chiropod* or Prosthetist*" or "Prosthetic*" or "Orthotist*" or "Orthotic*" or "Radiograph*" or "Speech and Language Therap*" or "speech therap*" or "SLT" or "Speech and language patholog*").ti,ab,kw,kf. **OR** exp Allied Health Personnel/ or exp Art Therapy/ or exp Nutritionists/ or exp Music Therapy/ or exp Occupational Therapy/ or exp Operating room technicians/ or exp Orthoptics/ or Osteopathic Manipulation/ or exp paramedics/ or exp Podiatry/ or "Prostheses and Implants"/ or Orthotic Devices/ or exp Speech-Language Pathology/ or exp Physical Therapists/ or exp emergency medical technicians/ or exp audiologists/ or physical therapy speciality/

**Concept:**

**2**	("Research*" adj (engag* or enhance or uptak* or capacity* or capabil* or participat* or cultur* or activ* or strateg*)).ti,ab,kw,kf.

**3**	Exp Research Personnel/

**4	2 or 3**

**Context UK:**

**5**	Exp United Kingdom/

**6**	(national health service* or nhs*).ti,ab,in.

**7**	(gb or "g.b." or (british* not "british columbia") or uk or "u.k." or united kingdom* or (england* not "new england") or northern ireland* or northern irish* or scotland* or scottish* or ((wales or "south wales") not "new south wales") or welsh*).ti,ab,jw,in.

**8**	(bath or "bath's" or (birmingham not alabama*) or ("birmingham's" not alabama*) or bradford or "bradford's" or brighton or "brighton's" or bristol or "bristol's" or carlisle* or "carlisle's" or (cambridge not (massachusetts* or boston* or harvard*)) or ("cambridge's" not (massachusetts* or boston* or harvard*)) or (canterbury not zealand*) or ("canterbury's" not zealand*) or chelmsford or "chelmsford's" or chester or "chester's" or chichester or "chichester's" or coventry or "coventry's" or derby or "derby's" or (durham not (carolina* or nc)) or ("durham's" not (carolina* or nc)) or ely or "ely's" or exeter or "exeter's" or gloucester or "gloucester's" or hereford or "hereford's" or hull or "hull's" or lancaster or "lancaster's" or leeds* or leicester or "leicester's" or (lincoln not nebraska*) or ("lincoln's" not nebraska*) or (liverpool not (new south wales* or nsw)) or ("liverpool's" not (new south wales* or nsw)) or (london not (ontario* or ont or toronto*)) or ("london's" not (ontario* or ont or toronto*)) or manchester or "manchester's" or (newcastle not (new south wales* or nsw)) or ("newcastle's" not (new south wales* or nsw)) or norwich or "norwich's" or nottingham or "nottingham's" or oxford or "oxford's" or peterborough or "peterborough's" or plymouth or "plymouth's" or portsmouth or "portsmouth's" or preston or "preston's" or ripon or "ripon's" or salford or "salford's" or salisbury or "salisbury's" or sheffield or "sheffield's" or southampton or "southampton's" or st albans or stoke or "stoke's" or sunderland or "sunderland's" or truro or "truro's" or wakefield or "wakefield's" or wells or westminster or "westminster's" or winchester or "winchester's" or wolverhampton or "wolverhampton's" or (worcester not (massachusetts* or boston* or harvard*)) or ("worcester's" not (massachusetts* or boston* or harvard*)) or (york not ("new york* or ny or ontario* or ont or toronto*)) or (york's not (new york*" or ny or ontario* or ont or toronto*))).ab,in,ti.

**9**	(bangor or "bangor's" or cardiff or "cardiffs" or newport or "newport's" or st asaph or "st asaph's" or st davids or swansea or "swansea's").ti,ab,in.

**10**	(aberdeen or "aberdeen's" or dundee or "dundee's" or edinburgh or "edinburgh's" or glasgow or "glasgow's" or inverness or (perth not australia*) or ("perth's" not australia*) or stirling or "stirling's").ti,ab,in.

**11**	(armagh or "armagh's" or belfast or "belfast's" or lisburn or "lisburn's" or londonderry or "londonderry's" or derry or "derry's" or newry or "newry's").ti,ab,in.

**12**	(exp africa/ or exp americas/ or exp antarctic regions/ or exp arctic regions/ or exp asia/ or exp oceania/) not (exp great britain/ or europe/)

**13	5 or 6 or 7 or 8 or 9 or 10 or 11**

**14	13 not 12**

**15	1 and 4 and 14**

**Search strategy- EMBASE via OVID**

**Population**

**1**	("Nurs*").ti,ab. **OR** exp nurse/ OR exp nursing/ OR nursing staff/ **OR** ("Midwife" OR "Midwives" OR "Midwif*").ti,ab. **OR** exp midwife/ or ("Allied Health*" OR "AHP*" OR "Professionals allied to health" OR "Art Therap*" OR "dietitian*" OR "dietetic*" OR "Drama therap*" OR "Music Therap*" OR "Occupational Therapist*" OR "OT*" OR "Operating Department Practitioner*" OR "ODP" OR "Orthoptist*" OR "Orthoptic*" OR "Osteopath*" OR "Paramedic*" OR "Podiatrist*" OR "Podiatry" OR "Chiropod*" OR "Prosthetist*" OR "Prosthetic*" OR "Orthotist*" OR "Orthotic*" OR "Radiograph*" OR "Speech and Language Therapist*" OR "SLT" OR "Speech and language patholog*").ti,ab. **OR** exp art therapy/ OR exp dietitian/ or exp drama therapy/ OR exp music therapy/ OR exp occupational therapy/ OR exp operating room personnel/ OR exp osteopathic medicine/ Or exp orthotist/ OR exp paramedical personnel/ OR exp prosthetist/ OR exp physiotherapist/ OR exp radiographer/ OR exp speech language pathologist/

**Concept**

**2**	("Research*" adj ("engag*" OR "enhance" OR "uptak*" OR "capacity*" OR "capabil*" OR "participat*" OR "outcome*" OR "cultur*" OR "activ*" OR "strateg*")).ti,ab. **OR** nursing research/

**Context UK**

**3**	exp United Kingdom/

**4**	(national health service* or nhs*).ti,ab,in,ad.

**5**	(English not ((published or publication* or translat* or written or language* or speak* or literature or citation*)adj5 english)).ti,ab.

**6**	(gb or "g.b." or (british* not "british columbia") or uk or "u.k." or united kingdom* or (england* not "new england") or northern ireland* or northern irish* or scotland* or scottish* or ((wales or "south wales") not "new south wales") or welsh*).ti,ab,jx,in,ad.

**7**	(bath or "bath's" or ((birmingham not alabama*) or ("birmingham's" not alabama*) or bradford or "bradford's" or brighton or "brighton's" or bristol or "bristol's" or carlisle* or "carlisle's" or (cambridge not (massachusetts* or boston* or harvard*)) or ("cambridge's" not (massachusetts* or boston* or harvard*)) or (canterbury not zealand*) or ("canterbury's" not zealand*) or chelmsford or "chelmsford's" or chester or "chester's" or chichester or "chichester's" or coventry or "coventry's" or derby or "derby's" or (durham not (carolina* or nc)) or ("durham's" not (carolina* or nc)) or ely or "ely's" or exeter or "exeter's" or gloucester or "gloucester's" or hereford or "hereford's" or hull or "hull's" or lancaster or "lancaster's" or leeds* or leicester or "leicester's" or (lincoln not nebraska*) or ("lincoln's" not nebraska*) or (liverpool not (new south wales* or nsw)) or ("liverpool's" not (new south wales* or nsw)) or ((london not (ontario* or ont or toronto*)) or ("london's" not (ontario* or ont or toronto*)) or manchester or "manchester's" or (newcastle not (new south wales* or nsw)) or ("newcastle's" not (new south wales* or nsw)) or norwich or "norwich's" or nottingham or "nottingham's" or oxford or "oxford's" or peterborough or "peterborough's" or plymouth or "plymouth's" or portsmouth or "portsmouth's" or preston or "preston's" or ripon or "ripon's" or salford or "salford's" or salisbury or "salisbury's" or sheffield or "sheffield's" or southampton or "southampton's" or st albans or stoke or "stoke's" or sunderland or "sunderland's" or truro or "truro's" or wakefield or "wakefield's" or wells or westminster or "westminster's" or winchester or "winchester's" or wolverhampton or "wolverhampton's" or (worcester not (massachusetts* or boston* or harvard*)) or ("worcester's" not (massachusetts* or boston* or harvard*)) or (york not ("new york*" or ny or ontario* or ont or toronto*)) or ("york's" not ("new york*" or ny or ontario* or ont or toronto*))))).ti,ab,in,ad.

**8**	(bangor or "bangor's" or cardiff or "cardiffs" or newport or "newport's" or st asaph or "st asaph's" or st davids or swansea or "swansea's").ti,ab,in,ad.

**9**	(aberdeen or "aberdeen's" or dundee or "dundee's" or edinburgh or "edinburgh's" or glasgow or "glasgow's" or inverness or (perth not australia*) or ("perth's" not australia*) or stirling or "stirling's").ti,ab,in,ad.

**10**	(armagh or "armagh's" or belfast or "belfast's" or lisburn or "lisburn's" or londonderry or "londonderry's" or derry or "derry's" or newry or "newry's").ti,ab,in,ad.

**11**	(exp “arctic and Antarctic”/ or exp oceanic regions/ or exp western hemisphere/ or exp africa/ or exp asia/)

**12**	(exp united kingdom/or Europe/)

**13**	**11 not 12**

**14**	**3 or 4 or 5 or 6 or 7 or 8 or 9 or 10**

**15**	**14 not 13**

**16**	**1 and 2 and 15**

**Search Strategy- CINAHL**

**Population**

**S1** Ti ("Nurse" OR "Nurses" OR "Nursing" OR "Nurs*") **OR** AB(("Nurse" OR "Nurses" OR "Nursing" OR "Nurs*")) **OR** (MH "Nurses" OR MH "Nursing" OR MH "Nursing Staff" OR MH "Practical Nursing" OR MH "Nursing Care")

**S2** TI("Midwife" OR "Midwives" OR "Midwif*")  **OR** AB ("Midwife" OR "Midwives" OR "Midwif*") **OR** (MH "Midwifery")

**S3** Ti ("Allied Health*" OR "AHP*" OR "Professionals allied to health" OR "Art Therap*" OR "dietitian*" OR "dietetic*" OR "Dramatherap*" OR "Drama Therapy" OR "Music Therapist" OR "Music Therap*" OR "Occupational Therapist*" OR "OT*" OR "Operating Department Practitioner*" OR "ODP" OR "Orthoptist*" OR "Orthoptic*" OR "Osteopath*" OR "Paramedic*" OR "Podiatrist*" OR "Podiatry" OR "Chiropod*" OR "Prosthetist*" OR "Prosthetic*" OR "Orthotist*" OR "Orthotic*" OR "Radiograph*" OR "Speech and Language Therapist*" OR "SLT" OR "Speech and language patholog*") **OR** AB ("Allied Health*" OR "AHP*" OR "Professionals allied to health" OR "Art Therap*" OR "dietitian*" OR "dietetic*" OR "Dramatherap*" OR "Drama Therapy" OR "Music Therapist" OR "Music Therap*" OR "Occupational Therapist*" OR "OT*" OR "Operating Department Practitioner*" OR "ODP" OR "Orthoptist*" OR "Orthoptic*" OR "Osteopath*" OR "Paramedic*" OR "Podiatrist*" OR "Podiatry" OR "Chiropod*" OR "Prosthetist*" OR "Prosthetic*" OR "Orthotist*" OR "Orthotic*" OR "Radiograph*" OR "Speech and Language Therapist*" OR "SLT" OR "Speech and language patholog*") **OR** (MH "Allied Health Personnel" OR MH "Art Therapy" OR MH "Dietitians" OR MH "Music Therapy" OR MH "Occupational Therapy" OR MH "Surgical Procedures, Operative" OR MH "Orthoptics" OR MH "Osteopathic Medicine" OR MH "Emergency Medical Technicians" OR MH "Podiatry" OR MH "Prostheses and Implants" OR MH "Orthotic Devices" OR MH "Radiologic Technologists" OR MH "Diagnostic Imaging" OR MH "Speech-Language Pathology" OR MH "Physical Therapists")

**Concept**

**S4** Ti ("Research*" N3 ("engag*" OR "enhance" OR "uptak*" OR "capacity*" OR "capabil*" OR "participat*" OR "outcome*" OR "cultur*" OR "activ*" OR "strateg*")) **OR** AB ("Research*" N0 ("engag*" OR "enhance" OR "uptak*" OR "capacity*" OR "capabil*" OR "participat*" OR "outcome*" OR "cultur*" OR "activ*" OR "strateg*")) **OR** (MH "Research Personnel")

**Context- UK**

**S5** (MH "United Kingdom+")

**S6** Ti (national health service* or nhs*) **OR** AB (national health service* or nhs*) **OR** AF (national health service* or nhs)

**S7** TI ( (english not ((published or publication* or translat* or written or language* or speak* or literature or citation*) N5 english) ) OR AB ( (english not ((published or publication* or translat* or written or language* or speak* or literature or citation*) N5 english) )

**S8** TI ( (gb or "g.b." or britain* or (british* not "british columbia") or uk or "u.k." or united kingdom* or (england* not "new england") or northern ireland* or northern irish* or scotland* or scottish* or ((wales or "south wales") not "new south wales") or welsh*) ) OR AB ( (gb or "g.b." or britain* or (british* not "british columbia") or uk or "u.k." or united kingdom* or (england* not "new england") or northern ireland* or northern irish* or scotland* or scottish* or ((wales or "south wales") not "new south wales") or welsh*) ) OR AF ( (gb or "g.b." or britain* or (british* not "british columbia") or uk or "u.k." or united kingdom* or (england* not "new england") or northern ireland* or northern irish* or scotland* or scottish* or ((wales or "south wales") not "new south wales") or welsh*) ) OR AB ((gb or "g.b." or (british* not "british columbia") or uk or "u.k." or united kingdom* or (england* not "new england") or northern ireland* or northern irish* or scotland* or scottish* or ((wales or "south wales") not "new south wales") or welsh*))

**S9** TI ( (bath or "bath's" or ((birmingham not alabama*) or ("birmingham's" not alabama*) or bradford or "bradford's" or brighton or "brighton's" or bristol or "bristol's" or carlisle* or "carlisle's" or (cambridge not (massachusetts* or boston* or harvard*)) or ("cambridge's" not (massachusetts* or boston* or harvard*)) or (canterbury not zealand*) or ("canterbury's" not zealand*) or chelmsford or "chelmsford's" or chester or "chester's" or chichester or "chichester's" or coventry or "coventry's" or derby or "derby's" or (durham not (carolina* or nc)) or ("durham's" not (carolina* or nc)) or ely or "ely's" or exeter or "exeter's" or gloucester or "gloucester's" or hereford or "hereford's" or hull or "hull's" or lancaster or "lancaster's" or leeds* or leicester or "leicester's" or (lincoln not nebraska*) or ("lincoln's" not nebraska*) or (liverpool not (new south wales* or nsw)) or ("liverpool's" not (new south wales* or nsw)) or ((london not (ontario* or ont or toronto*)) or ("london's" not (ontario* or ont or toronto*)) or manchester or "manchester's" or (newcastle not (new south wales* or nsw)) or ("newcastle's" not (new south wales* or nsw)) or norwich or "norwich's" or nottingham or "nottingham's" or oxford or "oxford's" or peterborough or "peterborough's" or plymouth or "plymouth's" or portsmouth or "portsmouth's" or preston or "preston's" or ripon or "ripon's" or salford or "salford's" or salisbury or "salisbury's" or sheffield or "sheffield's" or southampton or "southampton's" or st albans or stoke or "stoke's" or sunderland or "sunderland's" or truro or "truro's" or wakefield or "wakefield's" or wells or westminster or "westminster's" or winchester or "winchester's" or wolverhampton or "wolverhampton's" or (worcester not (massachusetts* or boston* or harvard*)) or ("worcester's" not (massachusetts* or boston* or harvard*)) or (york not ("new york*" or ny or ontario* or ont or toronto*)) or ("york's" not ("new york*" or ny or ontario* or ont or toronto*))))) ) OR AB ( (bath or "bath's" or ((birmingham not alabama*) or ("birmingham's" not alabama*) or bradford or "bradford's" or brighton or "brighton's" or bristol or "bristol's" or carlisle* or "carlisle's" or (cambridge not (massachusetts* or boston* or harvard*)) or ("cambridge's" not (massachusetts* or boston* or harvard*)) or (canterbury not zealand*) or ("canterbury's" not zealand*) or chelmsford or "chelmsford's" or chester or "chester's" or chichester or "chichester's" or coventry or "coventry's" or derby or "derby's" or (durham not (carolina* or nc)) or ("durham's" not (carolina* or nc)) or ely or "ely's" or exeter or "exeter's" or gloucester or "gloucester's" or hereford or "hereford's" or hull or "hull's" or lancaster or "lancaster's" or leeds* or leicester or "leicester's" or (lincoln not nebraska*) or ("lincoln's" not nebraska*) or (liverpool not (new south wales* or nsw)) or ("liverpool's" not (new south wales* or nsw)) or ((london not (ontario* or ont or toronto*)) or ("london's" not (ontario* or ont or toronto*)) or manchester or "manchester's" or (newcastle not (new south wales* or nsw)) or ("newcastle's" not (new south wales* or nsw)) or norwich or "norwich's" or nottingham or "nottingham's" or oxford or "oxford's" or peterborough or "peterborough's" or plymouth or "plymouth's" or portsmouth or "portsmouth's" or preston or "preston's" or ripon or "ripon's" or salford or "salford's" or salisbury or "salisbury's" or sheffield or "sheffield's" or southampton or "southampton's" or st albans or stoke or "stoke's" or sunderland or "sunderland's" or truro or "truro's" or wakefield or "wakefield's" or wells or westminster or "westminster's" or winchester or "winchester's" or wolverhampton or "wolverhampton's" or (worcester not (massachusetts* or boston* or harvard*)) or ("worcester's" not (massachusetts* or boston* or harvard*)) or (york not ("new york*" or ny or ontario* or ont or toronto*)) or ("york's" not ("new york*" or ny or ontario* or ont or toronto*))))) ) OR AF ( (bath or "bath's" or ((birmingham not alabama*) or ("birmingham's" not alabama*) or bradford or "bradford's" or brighton or "brighton's" or bristol or "bristol's" or carlisle* or "carlisle's" or (cambridge not (massachusetts* or boston* or harvard*)) or ("cambridge's" not (massachusetts* or boston* or harvard*)) or (canterbury not zealand*) or ("canterbury's" not zealand*) or chelmsford or "chelmsford's" or chester or "chester's" or chichester or "chichester's" or coventry or "coventry's" or derby or "derby's" or (durham not (carolina* or nc)) or ("durham's" not (carolina* or nc)) or ely or "ely's" or exeter or "exeter's" or gloucester or "gloucester's" or hereford or "hereford's" or hull or "hull's" or lancaster or "lancaster's" or leeds* or leicester or "leicester's" or (lincoln not nebraska*) or ("lincoln's" not nebraska*) or (liverpool not (new south wales* or nsw)) or ("liverpool's" not (new south wales* or nsw)) or ((london not (ontario* or ont or toronto*)) or ("london's" not (ontario* or ont or toronto*)) or manchester or "manchester's" or (newcastle not (new south wales* or nsw)) or ("newcastle's" not (new south wales* or nsw)) or norwich or "norwich's" or nottingham or "nottingham's" or oxford or "oxford's" or peterborough or "peterborough's" or plymouth or "plymouth's" or portsmouth or "portsmouth's" or preston or "preston's" or ripon or "ripon's" or salford or "salford's" or salisbury or "salisbury's" or sheffield or "sheffield's" or southampton or "southampton's" or st albans or stoke or "stoke's" or sunderland or "sunderland's" or truro or "truro's" or wakefield or "wakefield's" or wells or westminster or "westminster's" or winchester or "winchester's" or wolverhampton or "wolverhampton's" or (worcester not (massachusetts* or boston* or harvard*)) or ("worcester's" not (massachusetts* or boston* or harvard*)) or (york not ("new york*" or ny or ontario* or ont or toronto*)) or ("york's" not ("new york*" or ny or ontario* or ont or toronto*))))) )

**S10** TI ( (bangor or "bangor's" or cardiff or "cardiff's" or newport or "newport's" or st asaph or "st asaph's" or st davids or swansea or "swansea's") ) OR AB ( (bangor or "bangor's" or cardiff or "cardiff's" or newport or "newport's" or st asaph or "st asaph's" or st davids or swansea or "swansea's") ) OR AF ( (bangor or "bangor's" or cardiff or "cardiff's" or newport or "newport's" or st asaph or "st asaph's" or st davids or swansea or "swansea's") ) **OR** TI ( (aberdeen or "aberdeen's" or dundee or "dundee's" or edinburgh or "edinburgh's" or glasgow or "glasgow's" or inverness or (perth not australia*) or ("perth's" not australia*) or stirling or "stirling's" ) OR AB ( (aberdeen or "aberdeen's" or dundee or "dundee's" or edinburgh or "edinburgh's" or glasgow or "glasgow's" or inverness or (perth not australia*) or ("perth's" not australia*) or stirling or "stirling's" ) OR AF ( (aberdeen or "aberdeen's" or dundee or "dundee's" or edinburgh or "edinburgh's" or glasgow or "glasgow's" or inverness or (perth not australia*) or ("perth's" not australia*) or stirling or "stirling's" )

**S11 S5 OR S6 OR S7 OR S8 OR S9 OR S10**

**S12** ((MH "Americas+")

**S13**(MH "Africa+")

**S14** (MH "Antarctic Regions")

**S15** (MH "Arctic Regions")

**S16** (MH "Asia+")

**S17** (MH "Indian Ocean Islands+")

**S18** (MH "Pacific Islands+")

**S19** (MH "Australia+")

**S20** (MH "Atlantic Islands+"))

**S21 S12 OR S13 OR S14 OR S15 or S16 or S17 or S18 or S19 or S20**

**S22** (MH "United Kingdom+") or (MH "Europe+")

**S23 S21 not S22**

**S24 S11 not S23**

**S25 S1 and S2 and S24**

**EMCARE – via OVID**

**Population**

**1**	("Nurs*").ti,ab. **OR** exp nurse/ OR exp nursing/ OR nursing staff/ **OR** ("Midwife" OR "Midwives" OR "Midwif*").ti,ab. **OR** exp midwife/ or ("Allied Health*" OR "AHP*" OR "Professionals allied to health" OR "Art Therap*" OR "dietitian*" OR "dietetic*" OR "Drama therap*" OR "Music Therap*" OR "Occupational Therapist*" OR "OT*" OR "Operating Department Practitioner*" OR "ODP" OR "Orthoptist*" OR "Orthoptic*" OR "Osteopath*" OR "Paramedic*" OR "Podiatrist*" OR "Podiatry" OR "Chiropod*" OR "Prosthetist*" OR "Prosthetic*" OR "Orthotist*" OR "Orthotic*" OR "Radiograph*" OR "Speech and Language Therapist*" OR "SLT" OR "Speech and language patholog*").ti,ab. **OR** exp art therapy/ OR exp dietitian/ or exp drama therapy/ OR exp music therapy/ OR exp occupational therapy/ OR exp operating room personnel/ OR exp osteopathic medicine/ Or exp orthotist/ OR exp paramedical personnel/ OR exp prosthetist/ OR exp physiotherapist/ OR exp radiographer/ OR exp speech language pathologist/

**Concept**

**2**	("Research*" adj ("engag*" OR "enhance" OR "uptak*" OR "capacity*" OR "capabil*" OR "participat*" OR "outcome*" OR "cultur*" OR "activ*" OR "strateg*")).ti,ab. **OR** nursing research/

**Context UK**

**3**	exp United Kingdom/

**4**	(national health service* or nhs*).ti,ab,in,ad.

**5**	(English not ((published or publication* or translat* or written or language* or speak* or literature or citation*)adj5 english)).ti,ab.

**6**	(gb or "g.b." or (british* not "british columbia") or uk or "u.k." or united kingdom* or (england* not "new england") or northern ireland* or northern irish* or scotland* or scottish* or ((wales or "south wales") not "new south wales") or welsh*).ti,ab,jx,in,ad.

**7**	(bath or "bath's" or ((birmingham not alabama*) or ("birmingham's" not alabama*) or bradford or "bradford's" or brighton or "brighton's" or bristol or "bristol's" or carlisle* or "carlisle's" or (cambridge not (massachusetts* or boston* or harvard*)) or ("cambridge's" not (massachusetts* or boston* or harvard*)) or (canterbury not zealand*) or ("canterbury's" not zealand*) or chelmsford or "chelmsford's" or chester or "chester's" or chichester or "chichester's" or coventry or "coventry's" or derby or "derby's" or (durham not (carolina* or nc)) or ("durham's" not (carolina* or nc)) or ely or "ely's" or exeter or "exeter's" or gloucester or "gloucester's" or hereford or "hereford's" or hull or "hull's" or lancaster or "lancaster's" or leeds* or leicester or "leicester's" or (lincoln not nebraska*) or ("lincoln's" not nebraska*) or (liverpool not (new south wales* or nsw)) or ("liverpool's" not (new south wales* or nsw)) or ((london not (ontario* or ont or toronto*)) or ("london's" not (ontario* or ont or toronto*)) or manchester or "manchester's" or (newcastle not (new south wales* or nsw)) or ("newcastle's" not (new south wales* or nsw)) or norwich or "norwich's" or nottingham or "nottingham's" or oxford or "oxford's" or peterborough or "peterborough's" or plymouth or "plymouth's" or portsmouth or "portsmouth's" or preston or "preston's" or ripon or "ripon's" or salford or "salford's" or salisbury or "salisbury's" or sheffield or "sheffield's" or southampton or "southampton's" or st albans or stoke or "stoke's" or sunderland or "sunderland's" or truro or "truro's" or wakefield or "wakefield's" or wells or westminster or "westminster's" or winchester or "winchester's" or wolverhampton or "wolverhampton's" or (worcester not (massachusetts* or boston* or harvard*)) or ("worcester's" not (massachusetts* or boston* or harvard*)) or (york not ("new york*" or ny or ontario* or ont or toronto*)) or ("york's" not ("new york*" or ny or ontario* or ont or toronto*))))).ti,ab,in,ad.

**8**	(bangor or "bangor's" or cardiff or "cardiffs" or newport or "newport's" or st asaph or "st asaph's" or st davids or swansea or "swansea's").ti,ab,in,ad.

**9**	(aberdeen or "aberdeen's" or dundee or "dundee's" or edinburgh or "edinburgh's" or glasgow or "glasgow's" or inverness or (perth not australia*) or ("perth's" not australia*) or stirling or "stirling's").ti,ab,in,ad.

**10**	(armagh or "armagh's" or belfast or "belfast's" or lisburn or "lisburn's" or londonderry or "londonderry's" or derry or "derry's" or newry or "newry's").ti,ab,in,ad.

**11**	(exp “arctic and Antarctic”/ or exp oceanic regions/ or exp western hemisphere/ or exp africa/ or exp asia/)

**12**	(exp united kingdom/or Europe/)

**13**	**11 not 12**

**14**	**3 or 4 or 5 or 6 or 7 or 8 or 9 or 10**

**15**	**14 not 13**

**16**	**1 and 2 and 15**

**BNI- Via ProQuest**

1 ("Nurs*")
2 MAINSUBJECT.EXACT("Nurses")
3 MAINSUBJECT.EXACT("Nursing")
4 ("Midwives" or "Midwif*")
5 MAINSUBJECT.EXACT("Midwifery")
6 **(**"Allied Health* " or "AHP*" or "Professional* allied to health*" or "Art Therap*" or "dietitian*" or "dietetic*" or "Drama therap*" or "Music Therap*" or "Occupational Therap*" or "OT**" or "Operating Department Practitioner*" or "Operating room technician*" or "ODP" or "Orthoptist*" or "Orthoptic*" or "Osteopath*" or "Paramedic*" or "Podiatrist*" or "Podiatry" or "Chiropod* or Prosthetist*" or "Prosthetic*" or "Orthotist*" or "Orthotic*" or "Radiograph*" or "Speech and Language Therap*" or "speech therap*" or "SLT" or "Speech and language patholog*")
7 MAINSUBJECT.EXACT("Art therapy")
8 MAINSUBJECT.EXACT("Dietitians")
9 MAINSUBJECT.EXACT("creative Therapy")
10 MAINSUBJECT.EXACT("Music therapy")
11 MAINSUBJECT.EXACT("Occupational therapy")
12 MAINSUBJECT.EXACT("Paramedics")
13 MAINSUBJECT.EXACT("Podiatry")
14 MAINSUBJECT.EXACT("Speech therapy")

**Concept:**

**2**	("Research*" adj (engag* or enhance or uptak* or capacity* or capabil* or participat* or cultur* or activ* or strateg*))

**Context UK:**

**6**	(national health service* or nhs*)

**7**	(gb or "g.b." or (british* not "british columbia") or uk or "u.k." or united kingdom* or (england* not "new england") or northern ireland* or northern irish* or scotland* or scottish* or ((wales or "south wales") not "new south wales") or welsh*)

**8**	(bath or "bath's" or (birmingham not alabama*) or ("birmingham's" not alabama*) or bradford or "bradford's" or brighton or "brighton's" or bristol or "bristol's" or carlisle* or "carlisle's" or (cambridge not (massachusetts* or boston* or harvard*)) or ("cambridge's" not (massachusetts* or boston* or harvard*)) or (canterbury not zealand*) or ("canterbury's" not zealand*) or chelmsford or "chelmsford's" or chester or "chester's" or chichester or "chichester's" or coventry or "coventry's" or derby or "derby's" or (durham not (carolina* or nc)) or ("durham's" not (carolina* or nc)) or ely or "ely's" or exeter or "exeter's" or gloucester or "gloucester's" or hereford or "hereford's" or hull or "hull's" or lancaster or "lancaster's" or leeds* or leicester or "leicester's" or (lincoln not nebraska*) or ("lincoln's" not nebraska*) or (liverpool not (new south wales* or nsw)) or ("liverpool's" not (new south wales* or nsw)) or (london not (ontario* or ont or toronto*)) or ("london's" not (ontario* or ont or toronto*)) or manchester or "manchester's" or (newcastle not (new south wales* or nsw)) or ("newcastle's" not (new south wales* or nsw)) or norwich or "norwich's" or nottingham or "nottingham's" or oxford or "oxford's" or peterborough or "peterborough's" or plymouth or "plymouth's" or portsmouth or "portsmouth's" or preston or "preston's" or ripon or "ripon's" or salford or "salford's" or salisbury or "salisbury's" or sheffield or "sheffield's" or southampton or "southampton's" or st albans or stoke or "stoke's" or sunderland or "sunderland's" or truro or "truro's" or wakefield or "wakefield's" or wells or westminster or "westminster's" or winchester or "winchester's" or wolverhampton or "wolverhampton's" or (worcester not (massachusetts* or boston* or harvard*)) or ("worcester's" not (massachusetts* or boston* or harvard*)) or (york not ("new york* or ny or ontario* or ont or toronto*)) or (york's not (new york*" or ny or ontario* or ont or toronto*))).ab,in,ti.

**9**	(bangor or "bangor's" or cardiff or "cardiffs" or newport or "newport's" or st asaph or "st asaph's" or st davids or swansea or "swansea's")

**10**	(aberdeen or "aberdeen's" or dundee or "dundee's"or edinburgh or "edinburgh's" or glasgow or "glasgow's" or inverness or (perth not australia*) or ("perth's" not australia*) or stirling or "stirling's")

**11**	(armagh or "armagh's" or belfast or "belfast's" or lisburn or "lisburn's" or londonderry or "londonderry's" or derry or "derry's" or newry or "newry's")

**Search Terms to be used for Stakeholders websites and Overton, Google first 100 results**

- Nurse
- Midwife
- AHP
- Allied Health Professional
- Success
- Aims
- Goals
- Research Capacity
- Research Capability
